# SARS-CoV-2 infection dynamics revealed by wastewater sequencing analysis and deconvolution

**DOI:** 10.1101/2021.11.30.21266952

**Authors:** Vic-Fabienne Schumann, Rafael Ricardo de Castro Cuadrat, Emanuel Wyler, Ricardo Wurmus, Aylina Deter, Claudia Quedenau, Jan Dohmen, Miriam Faxel, Tatiana Borodina, Alexander Blume, Martin Meixner, José Horacio Grau, Karsten Liere, Thomas Hackenbeck, Frederik Zietzschmann, Regina Gnirss, Uta Böckelmann, Bora Uyar, Vedran Franke, Niclas Barke, Janine Altmüller, Nikolaus Rajewsky, Markus Landthaler, Altuna Akalin

## Abstract

The use of RNA sequencing from wastewater samples is a valuable way for estimating infection dynamics and circulating lineages of SARS-CoV-2. This approach is independent from testing individuals and can therefore become the key tool to monitor this and potentially other viruses. However, it is equally important to develop easily accessible and scalable tools which can highlight critical changes in infection rates and dynamics over time across different locations given sequencing data from wastewater. Here, we provide an analysis of lineage dynamics in Berlin and New York City using wastewater sequencing and present PiGx SARS-CoV-2, a highly reproducible computational analysis pipeline with comprehensive reports. This end-to-end pipeline includes all steps from raw data to shareable reports, additional taxonomic analysis, deconvolution and geospatial time series analyses. Using simulated datasets (*in silico* generated and spiked-in samples) we could demonstrate the accuracy of our pipeline calculating proportions of Variants of Concern (VOC) from environmental as well as pre-mixed samples (spiked-in). By applying our pipeline on a dataset of wastewater samples from Berlin between February 2021 and January 2022, we could reconstruct the emergence of B.1.1.7(alpha) in February/March 2021 and the replacement dynamics from B.1.617.2 (delta) to BA.1 and BA.2 (omicron) during the winter of 2021/2022. Using data from very-short-reads generated in an industrial scale setting, we could see even higher accuracy in our deconvolution. Lastly, using a targeted sequencing dataset from New York City (receptor-binding-domain (RBD) only), we could reproduce the results recovering the proportions of the so-called cryptic lineages shown in the original study. Overall our study provides an in-depth analysis reconstructing virus lineage dynamics from wastewater, and that our tool can be used to identify new mutations and to detect any emerging new lineages with different amplification and sequencing methods. Our approach can support efforts to establish continuous monitoring and early-warning projects for detecting SARS-CoV-2 or any other pathogen.

## Introduction

The ongoing COVID-19 pandemic highlighted the need for monitoring approaches to track emerging pathogens and pathogenic lineages. Acknowledging the importance and potential impact of wastewater-borne epidemiological analysis, the European Commission has recently recommended to implement continuous monitoring on SARS-CoV-2 through wastewater in all member states [1]. SARS-CoV-2 is a positive strand RNA virus from the family Coronaviridae, genus *Betacoronavirus* [2, 3]. As an alternative to individual patient tests that are tedious and expensive, Wastewater Based Epidemiology (WBE) has, before this pandemic, been used for different enteric microorganisms such as vaccine and wildtype polioviruses [4], rotaviruses, hepatitis A, astroviruses, adenoviruses, and noroviruses [5]. In the past two years, wastewater monitoring has been shown to be an effective tool for monitoring incidence rates. Multiple studies showed that it is possible to detect viral RNA even before widespread clinical reports [6–9], suggesting a potential as an early alert system.

Several WBE initiatives for SARS-CoV-2 monitoring were established worldwide, and currently, the COVIDpoops19 initiative [10] lists 128 dashboards from 276 universities monitoring 3364 sites. However, many of those studies are based on RT-qPCR analyses, limited to quantifying the viral titer and/or tracking a few known lineages, correlating the results with the reported number of cases in the area. A few studies have been using amplicon sequencing or metagenomics covering the whole viral genome, allowing to track virus lineages through signature mutations [11–13]. However, quantifying Variants of Concern (VOC) by next generation sequencing (NGS) reads remains challenging due to fragmented sequences. Moreover, sequencing and quantifying lineages are just the first steps in understanding the dynamics of the outbreaks. The sequencing results should be easily analyzed and combined with geospatial time series analysis. Tracking of VOCs over time and space can inform policy-making decisions in order to control new outbreaks. In this study, we present a reproducible, open-source pipeline for analyzing continuous sampling of wastewater treatment plants to track signature mutations of SARS-CoV-2 lineages of interest and emerging mutations via wastewater amplicon sequencing. We first benchmarked the pipeline using simulated (*in silico*) data and spiked-in samples [14]. We also sequenced and analyzed samples from Berlin wastewater using the ARTIC protocol [15] with 2 different sequencing protocols of ∼250bp length (in the following called “dataset-Berlin250”) and under industry conditions of ∼35bp length (in the following called dataset-Berlin35) during the 3ed and 4th pandemic wave in Germany. Additionally, we analyzed previously published dataset from New York City, where the sequencing was restricted to the receptor binding domain (RBD) region [16] (in the following called “dataset-NYC(RBD)”), showing the accuracy and usefulness of our methods for SARS-CoV-2 monitoring with data generated from multiple sites and approaches (iSeq and MiSeq).

## Results

### A reproducible computational pipeline for tracking SARS-CoV-2 in wastewater

We developed a new pipeline - PiGx SARS-CoV-2 - in the framework of our previously published set of pipelines called PiGx [17]. They are designed with a special focus on usability and reproducibility. The new pipeline was added to the PiGx collection of pipelines and it is distributed together, using GNU Guix (See Figure 1 for a diagram of the workflow). The pipeline comes with all the needed tools and their dependencies and can thus be reproduced on different systems independent of any other installed software.

**Figure 1:**
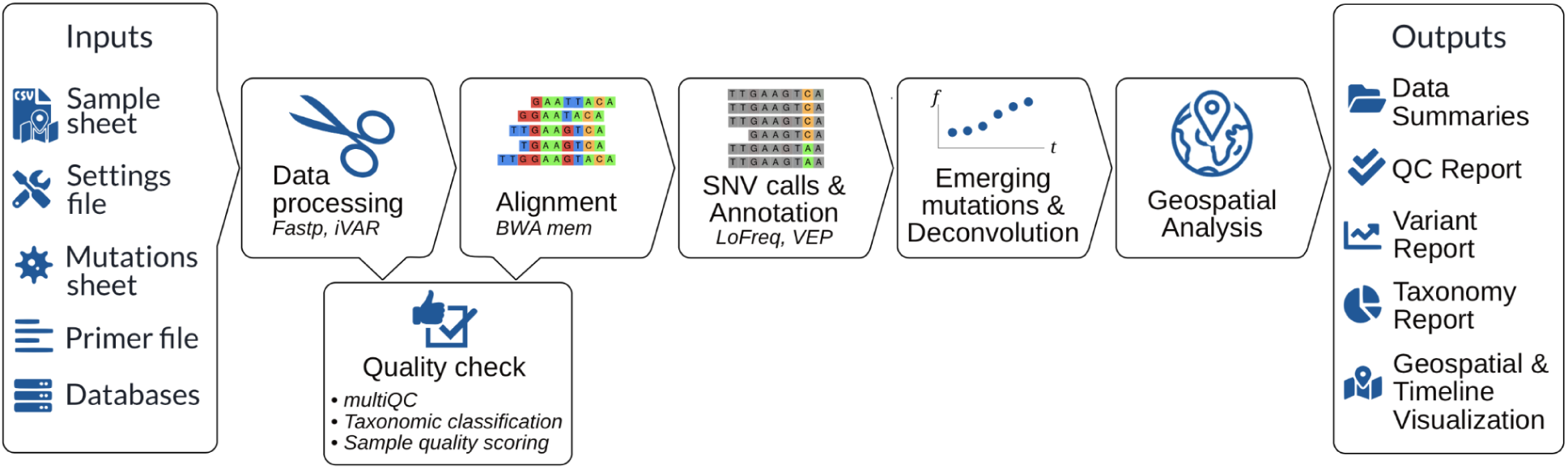
Flowchart of PiGx SARS-CoV-2 pipeline describing required input files, the analysis workflow and used tools and output files.

#### General description of the PiGx SARS-CoV-2 pipeline

The PiGx SARS-CoV-2 pipeline provides end-to-end data processing and analysis for wastewater RNA sequencing. The pipeline starts with a set of raw fastq read files, metadata such as locations and information about the lineages that should be tracked. After quality check and alignment, the variants are called and annotated. The samples from different timepoints are used to produce time-series reports that track trending mutations over time. We use a particular deconvolution step to also track the proportions of lineages representing Variants of Concern over time. Overall, the pipeline returns reports that provide overviews over lineage and single-mutation abundance in each sample, a taxonomic classification analysis of unaligned reads, and detailed quality control information. Furthermore, all per-sample results are summarized as tables and also combined to visualize time-series and geo-location plots, making the pipeline suitable for continuous sampling.

The pipeline needs local databases (downloaded by the user) for some of the annotation and alignment tools, such as *Ensembl VEP, Kraken2*, and *Krona tools*, while the tools themselves are automatically installed. Furthermore, the user needs to provide: (i) a sample sheet (CSV format) containing information about sampling date and location; (ii) a settings file (YAML format) for specifying the experimental setup and optional custom parameter adjustments, (iii) a mutation sheet containing the lineages of interest and their signature mutations in nucleotide notation and and BED file containing their genomic coordinates; (iv) the reference genome of the target species (see Methods for a detailed description); (v) BED file containing the PCR primer locations (provided with the pipeline for ARTIC protocol).

To ensure reliable variant calling and robust lineage abundance prediction, the sample has to match stringent quality control measures. For this, information about the sequencing primers, adapters, and also a BED file containing the sites of the signature mutations is necessary. Specifically the latter is important to ensure comparability of the called variants across all processed samples.

Given these input files, the pipeline executes a series of quality check, alignment, variant calling, deconvolution and mutation trend analysis steps. In the end, it provides interactive reports with quality check, geospatial and time-series information for mutations and lineages, as well as downloadable files for the downstream analysis.

#### Benchmarking the pipeline using spiked-in and simulated samples

In order to check the accuracy of our pipeline, we analyzed two simulated datasets. First we analyzed a spike-in mixture dataset from Karthikeyan et al. [14]. We obtained 384 BAM files with reads pre-aligned to SARS-CoV-2 reference genome, with samples ranging from 1160 to 1,955,791 reads. Four samples did not pass the 90% reference genome coverage threshold and were discarded. On average, 99% of the signature mutation sites were covered with at least 100 reads. The mean number of signature mutations found per sample was 37 out of 99 tracked and the mean number of overall mutations was 225 (SD 54).

Analyzing the predictions for each lineage with our deconvolution method, we found that the prediction robustness varies between lineages (Figure 2A), but overall, we were able to recall the expected proportions of lineages with R^2^ ∼0.74 (Figure 2B), excluding lineage A. In the samples with lineage A, only a maximum of 3 unique signature mutations for this lineage were found across all samples. Therefore, we got the weakest prediction for lineage A with an R^2^ of only 0.48. Because of this inconclusive result for lineage A, we decided to exclude its values from the general analysis mentioned above and shown in Figure 2B. The predictions for the other lineages show more accurate results (R^2^ between 0.71 - 0.86).

**Figure 2:**
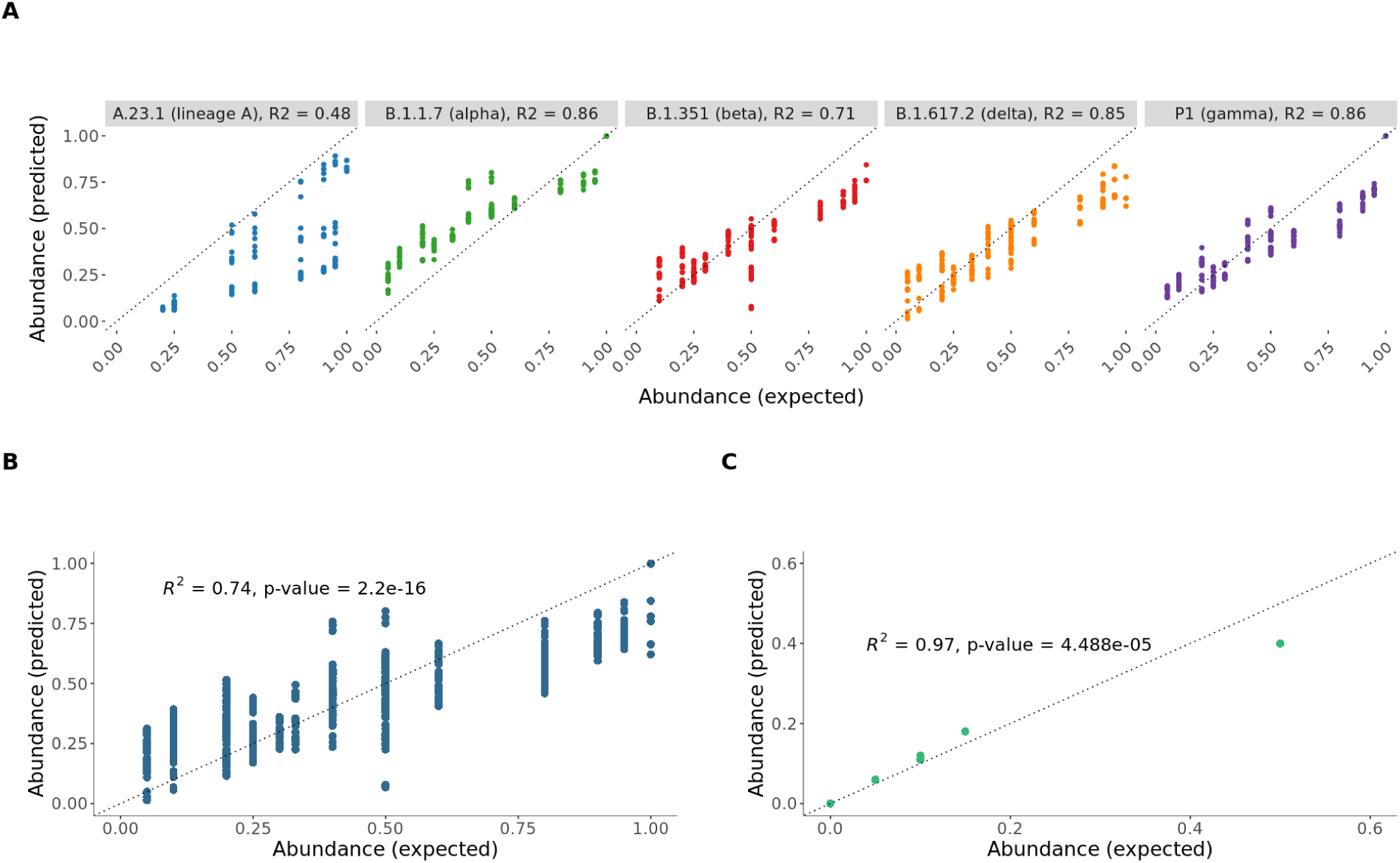
A) Prediction verification results for the spike-in data simulation per lineage; B) Prediction verification results for the spike-in data simulation across all lineages excluding lineage A; C) Prediction verification results *in-silico* simulation, single-end simulated 40bp reads from GISAID, 100k reads

Additionally, we tested the pipeline on a second simulation data set (see Methods), generated *in silico* with known proportions of lineages. A total of 100,000 reads were generated, and 98.4% of the reads were successfully aligned to the reference genome. All signature mutation genomic sites were covered with at least 10 reads and 74 of 179 signature mutations were found. Overall, our methods were able to get the proportions of lineages with an average error of 3% showing a maximum error of 10%. (Figure 2C).

### Wastewater SARS-CoV-2 sequencing and analysis with PiGx SARS-CoV-2

For this study, we sequenced a total of 988,025,456 reads from 171 samples from Berlin, using two different sequencing protocols. Firstly, for dataset-Berlin250 we obtained 74,633,648 reads, from 62 samples collected at four different wastewater treatment plants in Berlin operated by the municipal water authority (“Berliner Wasserbetriebe”) from 09^th^ of February to 10^th^ of June 2021 (Phase I) and from 16^th^ of September 2021 to 19^th^ of January 2022 (Phase II), using paired-end Miseq/Novaseq protocol with 2×250 bp reads. Between the two phases, due to low incidence rates, sequencing quality was insufficient. The average number of read-covered signature mutation sites per sample was 105 (SD 33, from a total of 154 tracked signature mutations, see mutation tables in the Supplementary Table S1). Of those 62 samples, 16 samples did not pass the defined quality control threshold (samples for which less than 90% of the signature mutation sites were covered).

We were able to align from 11 to 99 percent of sequencing reads (1 outlier with only 5% aligned reads) to the Wuhan reference SARS-CoV-2 genome, and the resulting alignments were used for variant calling. We were able to detect a total of 3,210 mutations, of which 133 are signature mutations, across all the samples (See methods for details on alignment and variant calling). The overall frequency of mutations per sample is shown on Supplementary Table S2.1-S2.4. The results of the time-series analysis for mutations and deconvolution of lineages for this dataset is presented in the sections below.

Secondly, industry scale dataset-Berlin35 contains 109 samples from one Berlin wastewater plant and three pumpstations (also operated by “Berliner Wasserbetriebe”). We used a paired-end very-short-read protocol (2 × 35 bp), for fast real time monitoring from 03.08.2021 to 20.01.2022. This data was analyzed in order to test our pipeline in a real time data monitoring system. We obtained a total of 913,391,808 35 bp reads. The average reference genome coverage was 97% (SD 5.7) with 9 samples not passing the quality control (QC) criteria of >90% reference genome coverage. The average number of signature mutations found per sample was 27 from the 154 tracked (SD 14.5) mutations and the mean of overall mutations found was 288 per sample (SD 147.5). The results of time-series mutation analysis and deconvolution of lineages for this dataset can be found in Supplementary Table S3.

The third dataset - dataset-NYC(RBD) - originated from published deep sequencing data of the receptor binding domain (RBD) of SARS-CoV-2 on samples from January 31^th^ to June 14^th^ 2021 collected in New York City (NYC) wastewater and published by Smyth et. al [16]. In the 94 samples reanalyzed here, we found, on average, 8 of the 12 mutation sites within the RBD (mean number of signature mutations found was 3.5). We did not apply a reference genome coverage cutoff because the sequencing was restricted to a small genomic region.

Smyth et. al described 3 cryptic lineages that where found in New York City wastewater: WWTF #10 (7 mutations), WWTF #11 (8 mutations), WWTF #3 (23 mutations) (see Supplementary Table S4). We tested our pipeline’s ability to highlight those “cryptic lineages” early on as well from the purely computational analysis in contrast to the extensive wet lab experiments that it took the authors to discover them. The results of this analysis are shown in the section below.

### Emerging mutations can be teased out from time-series analysis

The time-series nature of the data can not only be used to track SARS-CoV-2 lineages, but also to identify trends for individual mutations. We applied a linear regression model for each mutation using the date of sampling as the independent variable to identify mutations with strong increasing trends over time (see Methods). We considered mutations significant if the t-test p-value is below 0.05. The full lists can be found in Supplementary Table S5.

Overall, merging the two sample groups within dataset-Berlin250 from Berlin Phase I and II for a single analysis, 105 mutations were significantly changing over time from February 2021 until January 2022. The top 10 most significantly changed mutations are shown in Figure 3A.

**Figure 3:**
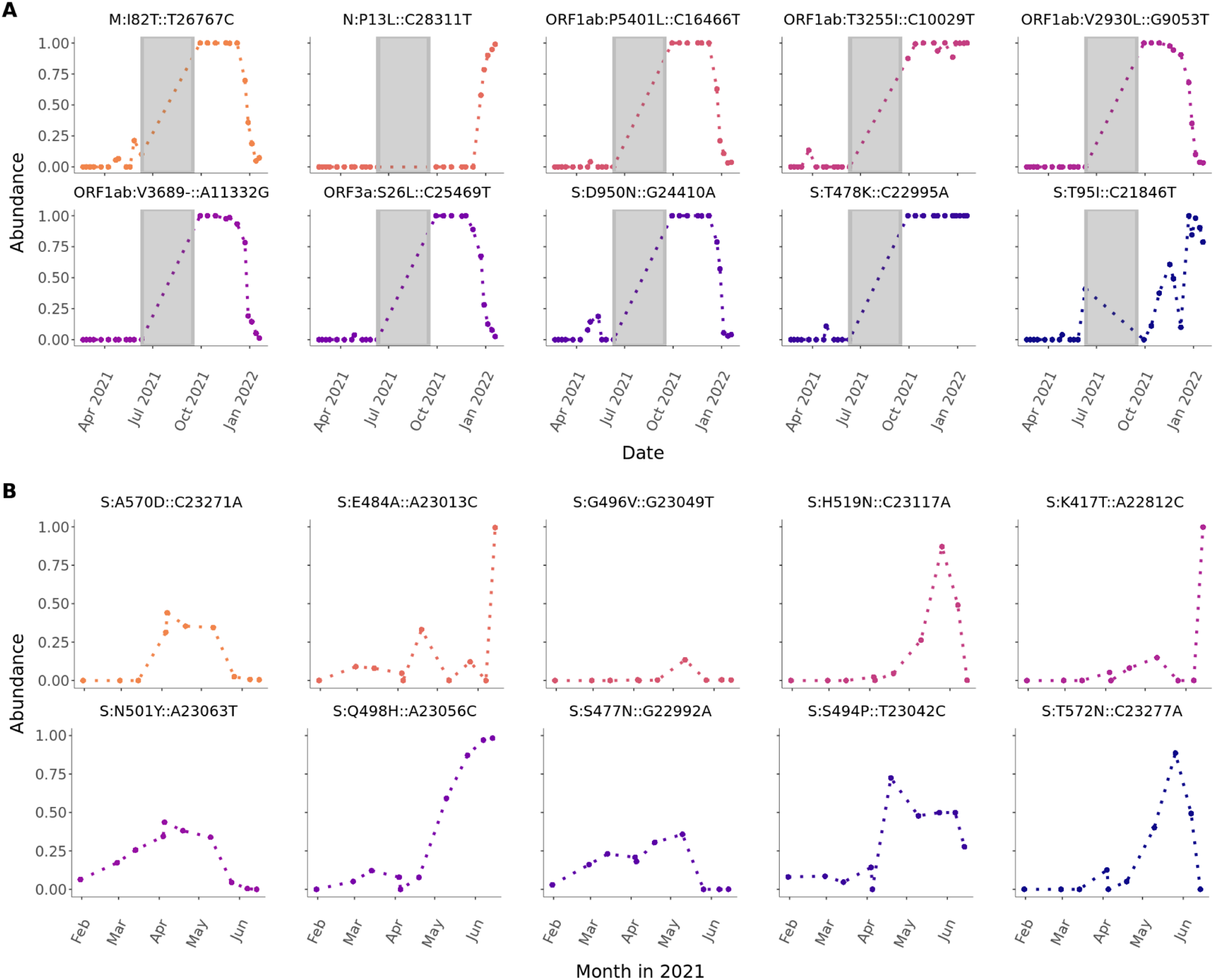
A) Top 10 sequence variants that significantly increase over time in Berlin. The mutations were pooled over locations of four different wastewater treatment plants and daytime and sorted by decreasing coefficients from linear models. Statistical significance was evaluated by a t-test using p <= 0.05 as cutoff. Only samples passing the sample quality scoring (> 90% reference genome coverage) were used. There was no sampling between June 11 and September 19 2021. B) Top 10 sequence variants that significantly increase over time in New York City (NYC) (2021). The mutations were pooled over locations of 14 different wastewater treatment plants in NYC and daytime and sorted by decreasing coefficients from linear models. Statistical significance was evaluated by a t-test using p <= 0.05 as cutoff.

Here, six of the highlighted mutations M:I82T::T26767C, ORF3a:S26L::C25469T, ORF1ab:V3689-::A11332G, ORF1ab:V2930L::G9053T, S:D950N::G24410A, ORF1ab:P5401L::C16466T, are uniquely characteristic for the lineage B.1.617.2 (delta). They show a similar pattern, emerging mostly during the summer of 2021 and decreasing in January 2022. Hereby, S:D950N::G24410A and M:I82T::T26767C already started to appear with increasing frequency in late April 2021, but inconsistently. The mutation S:T478K::C22995A is a shared mutation between the lineages B.1.617.2 (delta), BA.1 and BA.2 (omicron). It showed a consistent increase from July 2021 and reached 100% of presence until the end of our time-series. However, N:P13L::C28311T and S:T95I::C21846T are unique mutations for the BA.1 lineage where the latter already started to continuously increase in frequency starting in October 2021 which is a month earlier than the B.1.1.529 (omicron) lineage family was started to track by the RKI (Figure 4C).

**Figure 4:**
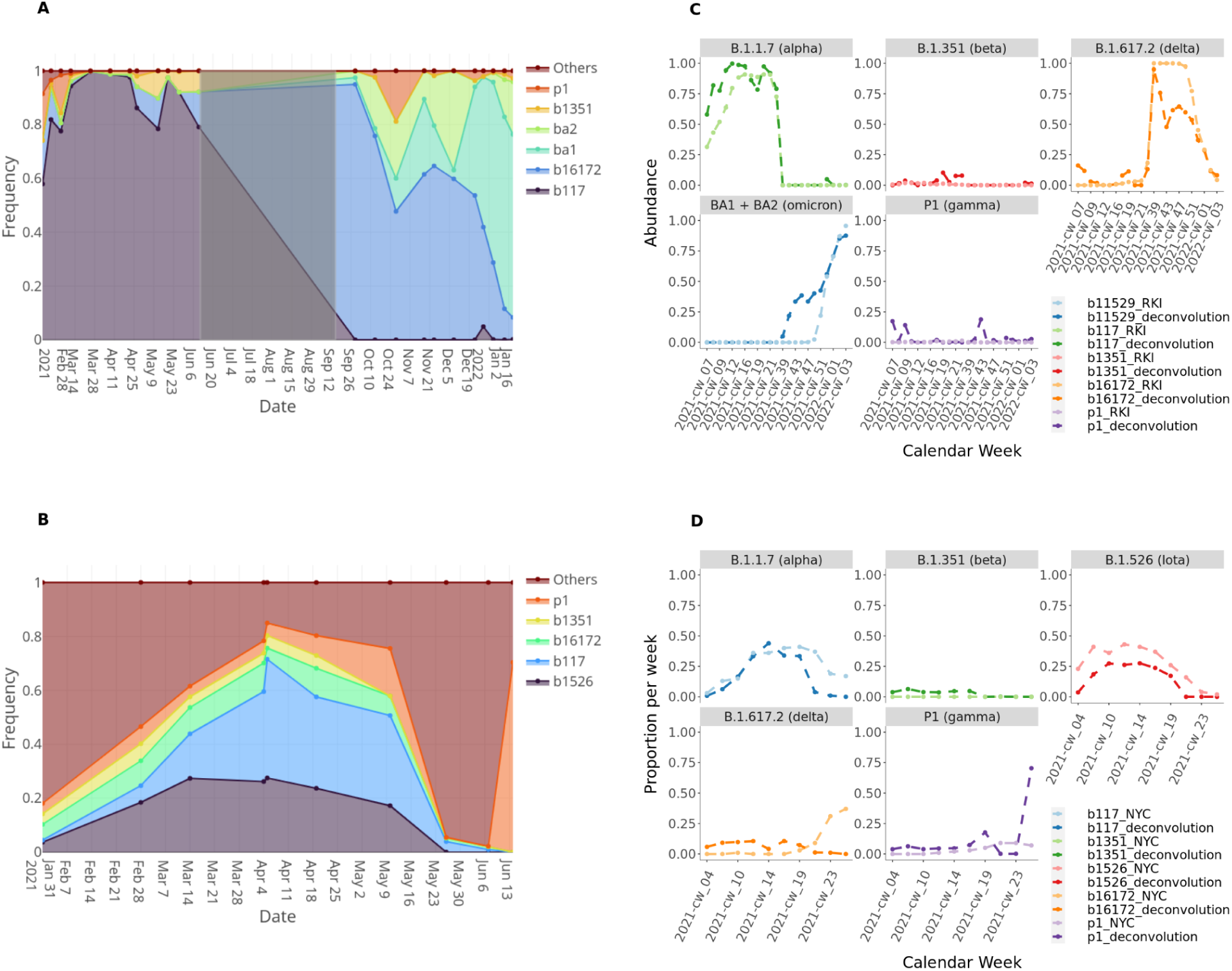
A) Proportion of tracked lineages over time in Berlin wastewater. Only samples passing the sample quality scoring (>=90% reference genome coverage) were considered. Shaded area highlights the non-sampling Phase. B) Proportion of tracked lineages over time in New York City wastewater. The proportions were calculated with a deconvolution model based on the signature mutation frequencies. “Others” denotes a set of reference mutations derived from the deconvolution matrix. Sample results were pooled from four different wastewater treatment plants using weighted mean with read number as weights. In case of undistinguishable lineages the proportion derived for the group was distributed equally for the affected lineages. C,D) Comparison of deconvolution results (dark color) with lineage frequency analysis data from Robert-Koch-Institute (RKI) (C) or NYC Department of Health and Mental Hygiene (NYC) (D) (light color). Deconvolution results were pooled by weeks using weighted mean using sample read numbers as weights. For the data from Berlin only samples passing the sample quality scoring (>=90% reference genome coverage) were used.

Within the dataset-NYC(RBD) [16], we found a total of 69 significantly changing genome variants. The highlighted mutations with the 10 highest correlation values in Figure 3B point out 8 of the 28 reported mutations of cryptic lineages: S:Q498H::A23056C, S:T572N::C23277A, S:H519N::C23117A, S:K417T::A22812C, S:E484A::A23013C, S:S494P::T23042C, S:S477N::G22992A. Additionally S:N501Y::A23063T and S:A570D::C23271A were highlighted which are characteristic mutations for B.1.1.7 (alpha). They show a constant increase already up to 40% in March which is around 1 month earlier than the reported abundancy for B.1.1.7 (alpha) based on cases (Figure 4D)

### Deconvolution of mutation frequencies infers SARS-CoV-2 lineage frequencies

In our pipeline, we have implemented methods to deconvolute the frequencies of VOCs from pooled sequencing reads. Briefly, the deconvolution method uses signature mutations for each VOC and tries to discern the proportions of these lineages making up the observed mutation frequencies in the pooled (bulk) sequencing reads obtained from the wastewater. In this study, we tracked 4 lineages which were classified at the time of data collection as VOCs: B.1.1.7 (alpha), B.1.351 (beta), P1 (gamma) and B.1.1617.2 (delta) in both datasets from Berlin and New York City. For the latter we additionally tracked the lineage B.1.526 (Iota). For the samples from Berlin from Phase II we additionally tracked the BA.1 and BA.2 lineages, which taxonomically are classified as sublineages of B.1.1.529 (omicron) and became VOCs in November 2021. We decided to track them separately in order to get a higher resolution on their dynamics. In the following, when comparing to official reported lineage abundances we are adding up our separate abundances for BA.1 and BA.2 to compare to reported values for B.1.1.529 (omicron). We characterized the lineages with a mutation table (Supplementary Table S1) containing signature nucleotide mutations from covidCG [18]. We took a list of mutations with a sequence consensus threshold of 70%. We included mutations that are unique for each lineage, as well as mutations that are shared by two or more lineages. Of note, the pipeline is flexible and can track any lineage if the signature mutations are provided in nucleotide format. We applied this deconvolution method (based on the frequencies of the signature mutations) to infer the proportions of each lineage on each sample (Supplementary Table S3). The lineage frequencies are predicted using a regression model based on the observed frequencies of the signature mutations for each lineage. Additionally, during the deconvolution process, we weighted the tracked lineages differently based on how many signature mutations were found for each of them for a given sample. This step is necessary in order to get more precise predictions of lineages with low abundance and for which only few or only shared mutations were found (see Methods).

Figure 4A shows VOC proportion changes over time across 4 wastewater treatment plants in Berlin (merged results of Phase I and Phase II). Overall, we predict an increase in B.1.1.7 (alpha) that had 57% on February 19^th^ (beginning of sampling of Phase I) and increased to 79% on June 10^th^ (end of sampling of Phase I) with a peak of 99% on May 25. Also B.1.351 (beta) increased from zero detection in February to 8% in May with a predicted peak of 10% on May 25. The B.1.617.2 (delta) lineage was barely detected with 3% over the sampling time of Phase I increasing to 11% on May 12. We predicted 16% of B.1.617.2 (delta) as early as in February 2021 but this result is likely to be inaccurate. For P1 we could predict in Phase I a decrease from 17 % on February 19 to zero in June. However in sampling Phase II, P1 is predicted again with an abundance peak of 18% on October 28. During winter 2021 the predicted P1 abundance decreases continuously down to 3% in January. The tracking of the lineages BA.1 and BA.2 started with sampling Phase II in September 2021 where they were initially predicted with a total abundance of 6%. Their abundance rapidly increased to ∼90% by January 19 with BA.1 at ∼70% and BA.2 with ∼20%. In the timeframe we sampled, the diversity and abundance of lineages that are not VOCs was already very reduced. We only predicted unspecified lineages (labeled as “Others”) with 8% in February 2021 and it fell below 1% on March 11 and never increased again.

In order to see if the predicted results can reflect the abundances of circulating lineages in Berlin, we compared the deconvolution results with lineage analysis data published by the Robert Koch-Institute (RKI) for Germany (Figure 4C). Hereby, lineage dynamics for Germany are very comparable to the dynamics within the city of Berlin. We can see that our predicted lineage frequencies are very similar to the reported lineage distribution based patient testing. Only B.1.1.7 (alpha) shows mostly higher predicted values, but with very similar trends. Also the predictions of the lineages BA.1 and BA.2, which are taken together comparable with the reported B.1.1.529 (omicron) values are higher in the beginning (December 2021) than the RKI values, but become very similar in January 2022. This is explainable with the continuous detection of the mutation ORF1ab:T3255I::C10029T, which is listed as unique signature mutation for BA.1 and BA.2, but is also carried by the B.1.617.2 sublineage 21J [19] (but not by the parent clade) that we did not actively track in this analysis.

The analysis of the dataset-Berlin35 showed similar results as for the dataset-Berlin250 as shown in Supplementary Figure 1. Of note, the prediction results for the abundances of B.1.617.2 (delta) and B.1.1.529/BA.1+BA.2 (omicron) are showing less divergence from the RKI values than for the dataset-Berlin250.

The data from New York City (Figure 4B) shows a more diverse mixture throughout the sampling phase according to the predicted high proportion of “Others”. This proportion was as high as 82% in January 2021 decreasing to 15% on April the 5^th^ but then had a predicted increase again to 97% in June. The most dominant lineages were B.1.1.7 (alpha) which increased from 0% in January up to 44% in April and B.1.526 (Iota) which increased from 4% in January up to predicted 28% in April. However, the comparison with the data reported from NYC Department of Health and Mental Hygiene (NYC health) (Figure 4D) suggests that both lineages circulated with similar abundances within the given timeframe and that the differences in the predicted values are due to the expected inaccuracy of the pipeline. The abundance of B.1.351 (beta) increased slightly from 4% in January to 5% in April but was not present anymore after. For B.1.617.2 (delta), we predicted a continued increase up to 10% in April which is also in contrast to the NYC health data where the abundance of delta only starts to increase at the end of May. For P1 (gamma) we predicted an increase from 3% in January up to 17% in May. This trend is also shown from the NYC health data. However, we also predicted 70% P1 in June. For this prediction only 4 signature mutations across all lineages were found and 1 of them is S:K417T::A22812C with a frequency of 1 which is a unique signature mutation of P1 (gamma). Besides both above-mentioned differences, our prediction results are consistent with the NYC health data as shown in Figure 4D. Unpooled results for single locations for both datasets are attached as Supplemental material (Supplementary Table S3).

In Figure 5, we combined the visualization of key mutation frequencies, cases of COVID-19 in Berlin (from RKI), and deconvolution results for B.1.617.2 (delta) and BA.1/BA.2 (omicron) lineages.

**Figure 5:**
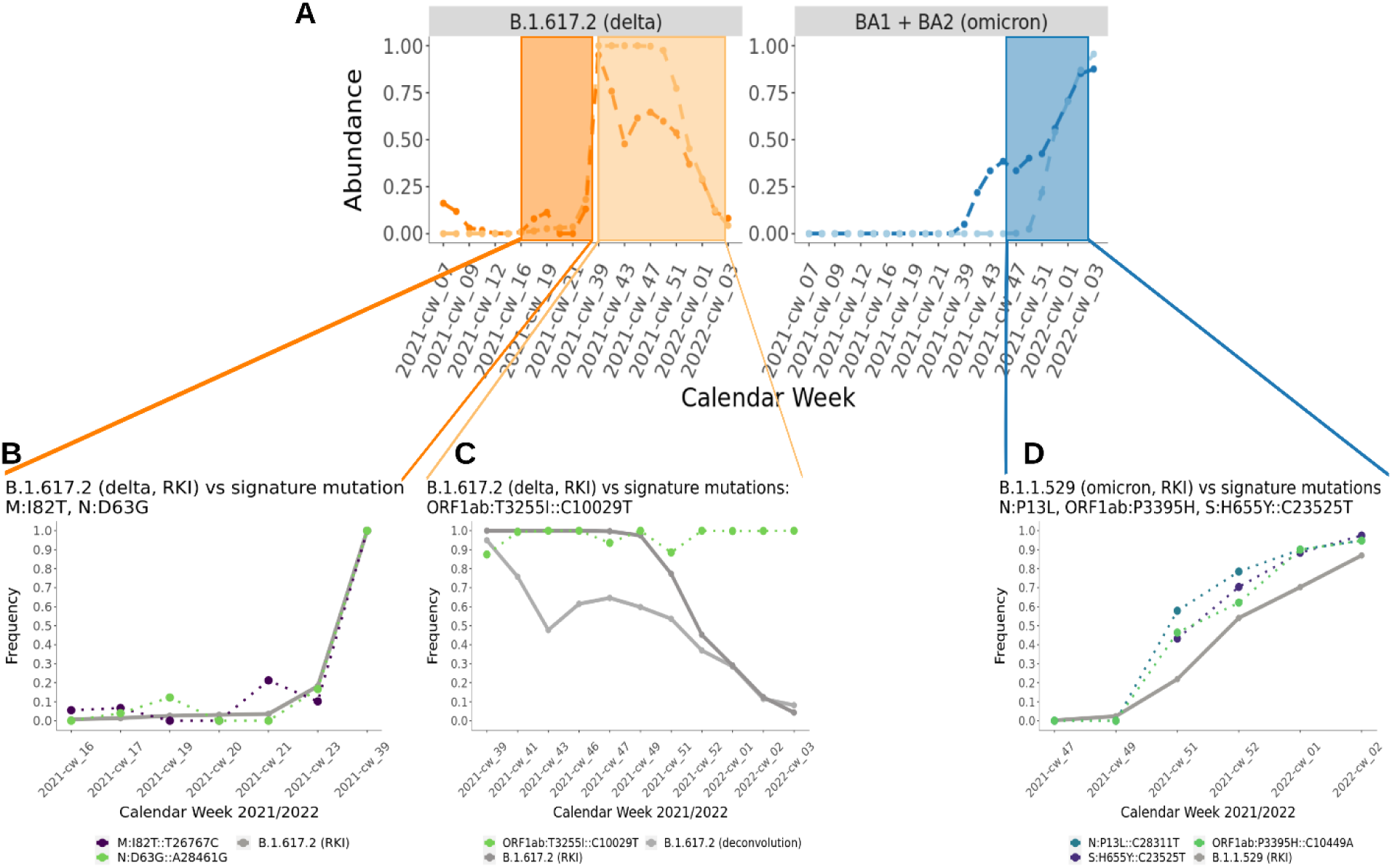
A) Combination of lineage prediction results (deconvolution) for B.1.617.2 and BA.1/BA.2 (dataset-Berlin250), B,C,D) single key signature mutations M:I82T::T26767C, N:D63G::A28461G, ORF1ab:T3255I::C10029T, ORF1ab:P3395H::C10449A, N:P13L::C28311T, S:H655Y::C23525T and case numbers in Berlin (from RKI).

We can see that the mutations M:I82T and M:D63G showed a strong increase together with RKI case numbers and with B.1.617.2 (delta) proportions from our deconvolution results. The same pattern is shown for N:P13L, ORF1ab:P3395H and S:H655Y, raising together with omicron lineages. However, the mutation ORF1ab:T3255I (tracked as BA.1 and BA.2 signature mutations in our deconvolution) was detected with frequency of 100% already in September 2021, while Omicron was not present yet. This mutation was in high frequency when B.1.617.2 (delta) was predominant and stayed high while omicron raised. This could have hinted that this mutation was already present in a sub-clade of B.1.617.2 (delta) [19] and in fact this mutation is present in delta sub-clade 21J [20].

### RT-qPCR on wastewater samples can predict SARS-CoV-2 rise 1-2 weeks in advance

We checked if RT-qPCR results were correlated with case numbers in the region. For RT-qPCR, we used 4 pairs of primers for SARS-CoV-2 detection (RT-qPCR) on the wastewater samples. Due to the very low amount of viral particles present overall, we decided for a semi-quantitative approach, instead of using the cycle threshold (Ct) values, calculating the number of positive detections divided by the number of total reactions carried, grouping all the samples for each day (See Methods for details). The daily percentage of positive qPCR reactions ranges from 0 to 83% (Supplementary Table S6). We also found positive, significant correlation with RT-qPCR results and incidence rates (adjusted R^2^ = 0.32, t-test p-value = 0.0004, see Figure 6A-B). In addition, we have also repeated the cross-correlation analysis between incidence rate and RT-qPCR results with different time lags. In this case, lag= -1 week also had positive correlation with the incidence rate (adjusted R^2^ = 0.47, coefficient = 0.5, t-test p-value = 9.8e-06) (Figure 6C-D). Additionally we checked the correlation between reference genome coverage and incidence cases in Berlin. The results did not show significant correlations between those variables (Supplementary Figure 3).

**Figure 6:**
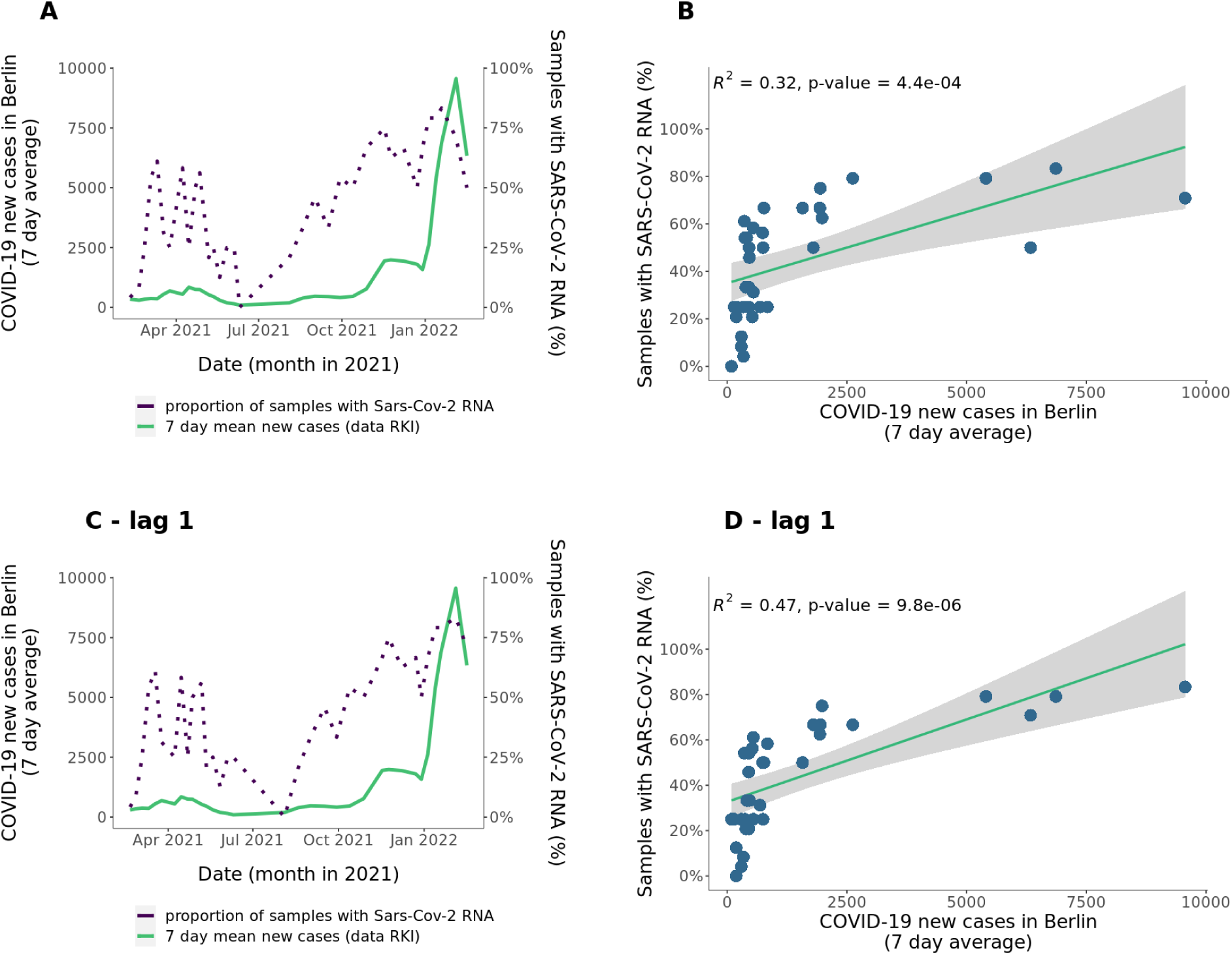
A) 7 days average of COVID-19 cases in Berlin, data from Robert Koch-Institute (RKI) (light green, left axis) and proportion of samples positively determined SARS-CoV-2 RNA by RT-qPCR (dark violet, right axis) over Feb - Jan 2022. B) Correlation of 7 days average of COVID-19 cases in Berlin and proportion of samples with positively determined SARS-CoV-2 RNA by RT-qPCR. C) 7 days average of COVID-19 cases in Berlin, data from Robert Koch-Institute (RKI) (light green, left axis) and proportion of samples positively determined SARS-CoV-2 RNA by RT-qPCR (dark violet, right axis) over Feb - Jan 2022 with one time point lag. D) Correlation of 7 days average of COVID-19 cases in Berlin and proportion of samples with positively determined SARS-CoV-2 RNA by RT-qPCR with one time point lag.

## Discussion

In many countries, epidemiological monitoring of SARS-CoV-2 is largely dependent on PCR-based or antigen detection methods without sequencing which is applied on patient samples. These techniques can be used for variant detection only after a concerning new lineage is detected and an appropriate assay was developed. In order to discover new lineages, we need to be able to call mutations of the SARS-CoV-2 genome which can be done using sequencing methods. However, sequencing-based techniques are deployed on only a fraction of the patient population. Wastewater monitoring emerged as a viable option to track the prevalence of COVID-19 and also for the emergence of different lineages [21] at the population level not only because it is faster and cheaper than sequencing of samples derived from patients, but it can also be more representative due to less bias through the choice of which samples are going to be sequenced. Furthermore it can also be used to track early emerging mutations or lineages of SARS-CoV-2. However, sequencing of SARS-CoV-2 material obtained from wastewater presents data analysis challenges as the samples are potentially from numerous patients, and have lower quality than material obtained directly from patients. In addition, the analytical workflows should be able to deal with samples from multiple locations and time points and combine the information in an easily accessible manner.

In order to address these challenges, we have built a reproducible analytics pipeline that takes in raw sequencing reads and provides sharable interactive reports with geospatial information, and mutation and lineage tracking features over time. In comparison to other commonly used pipelines for variant analysis like V-pipe [22] or the recommended ARTIC bioinformatics pipeline [23], PiGx SARS-CoV-2 additional features (discussed below) improved usability, reproducibility, and application for environmental samples like wastewater. In addition, the geospatial tracking allows to compare and monitor infection dynamics from different locations (See example reports in Data access section). In terms of usability, the novelty with PiGx SARS-CoV-2 is that the output reports include result visualization for each sample individually and also for the overview and summary of all samples with a choice of visualization methods that are straightforward to interpret. Furthermore, all outputs relevant for the assessment for lineages, quality control and mutations are produced in human-readable format such as HTML reports from which CSV files can be extracted. That makes further data analysis easier by providing formatted tables. Last but not least, PiGx SARS-CoV-2 offers state-of-the-art software reproducibility thanks to GNU Guix [17].

The pipeline comes with built-in flexible quality control metrics since samples from wastewater can have more frequent quality issues. In our analysis, we applied a strict cutoff for reference genome coverage (>=90%) for whole-genome sequencing data to reduce noise in our predictions. Our pipeline also allows the user to input their own reference genome and their own set of signature mutations and lineages. As an additional step for QC, we implemented a taxonomic classification of reads that did not align to the SARS-CoV-2 reference genome. Since we used a PCR based protocol, we expect some degree of nonspecific amplifications, so it is of great help to have an additional control by means of the taxonomic classification to assess potential biases. Also since *Kraken2* is a k-mer classifier, this method can reveal reads that match SARS-CoV-2 but are not aligned by stringent alignment tools. This is important to know because it provides insights about potential loss of new mutations missed on the alignment. This step allows the user to investigate potential issues and, if necessary, to adjust the alignment stringency.

Aiming to benchmark our pipeline, we tested it in two different simulated datasets, allowing us to estimate the error rates from our methods. For the dataset generated *in silico*, the difference between the expected and calculated frequency of lineages was only on average 3%. This shows that our pipeline has potential to track accurately VOC from sequencing data. However, we are aware that real sequencing data can offer further challenges, such as low quality sequencing, presence of many other microorganisms and untracked lineages of SARS-CoV-2. In order to better benchmark our tool, we also analyzed spiked-in samples generated by [14]. Our results overall performed worse than the original publication (our R^2^ for prediction of the mixture was ∼0.7 when the R^2^ from the original publication was 0.9). Those results can be explained by the differences in the mutations used for deconvolution in our analysis and the original study, highlighting the importance of carefully selecting and more importantly reporting the used mutation matrix.

One of the primary features of our approach is built-in tracking of emerging mutations. This feature allowed, for example, early prediction of lineages such as B.1.617.2 (delta) from a single signature mutation M:I82T::T26767C (Figure 5) in the dataset-Berlin250. We were able to detect the lineage characterizing mutations before the lineage itself was detected in the population (Figure 5). This specific mutation was described to be associated with critically increased viral fitness [24]. The analysis and results are also visualized without the need for any additional steps directly in the summarizing report. We showed that our pipeline and its reports can be a valuable tool for early warning predictions and to guide additional targeted analysis.

Another key feature of our approach is the deconvolution method that helps us identify the proportion of lineages present in environmental samples such as wastewater samples. By making use of a weighted regression method, we were able to provide accurate estimates of lineage proportions for our samples over time. For the VOCs that we tracked with signature mutations, we show in Figure 4 that our model can accurately predict the composition of lineages when comparing with abundances of circulating lineages reported during the same time frame, even with very low frequencies. This method was able to predict the rapid increase of the lineages BA.1 and BA.2 in the winter (Figure 4).

It is important to note that the mutations commonly used for tracking B.1.1.7 in other studies, S:N501Y::A23063T and del69/70 [25, 26] were rare or not found in our Berlin dataset, but they were detected in NYC dataset (Figure 3B), and this might be explained by PCR bias differences between the datasets, because the NYC dataset only sequenced the RBD genomic region, having a higher resolution on the mutations in this genomic region.

Additionally, with the dataset-Berlin35, we showed that our pipeline can be used in an industrial production system for real-time monitoring. The results obtained were comparable with the dataset-Berlin250 for the same time frame (Phase 2). Interestingly, for the dataset-Berlin35, BA.1+BA.2 (omicron) predictions are followed by RKI incidence cases closer on time than for the dataset-Berlin250, where we detect omicron and delta one week in advance. For B.1.617.2 (delta), the dataset-Berlin35 shows more similar proportions than for the dataset-Berlin250 (See Supplementary Figure 2). These results can suggest that the inaccuracies found in our dataset-Berlin250 can be explained by differences in the data generation (read length, internal sequencing validations or differences on sampling sites) rather than in data processing with our pipeline.

As reported in previous studies in other cities around the globe [27], we showed that also for Berlin, the quantification from wastewater can reveal infection dynamics potentially earlier than it is possible from clinical testing. Although RT-qPCR results are not fully quantitative, observing this expected trend was important and paved the way for more robust lineage and mutation trend analysis using sequencing.

Regardless of the methods used on wastewater, as previously published reports also indicate, wastewater monitoring may provide early warning for future case numbers and emerging mutations even post-pandemic when populations are not tested and monitored that thoroughly as during the pandemic.

In conclusion, we present a reproducible and comprehensive workflow with a strong emphasis on usability and reproducibility that has features for tracking mutations and VOC over time and geographical locations. We stress-tested the tool with simulated data and real world data from different locations and with different methods, showing the usefulness of our tool but also the importance of keeping lineage nomenclature and mutations tracked consistent, for comparable results.

## Methods

### Experimental methods

#### Enrichment of viral particles from raw wastewater and RNA extraction

For the dataset-Berlin250, raw wastewater samples were collected from four different wastewater treatment plants across Berlin, serving a population of approximately 3.4 million people in total. They were collected as composite two hour samples (8-10pm and 10-12pm) at the primary influent collector at the indicated wastewater treatment plants. Typical characteristics of Berlin wastewater treatment plant effluents are 500-1500 mg/L chemical oxygen demand, 200-600 mg/L suspended solids, 40-80 mg/L ammonium-N, 2-8 mg/L orthophosphate-P, 1500-2000 µS/cm electrical conductivity.

Samples were stored and transported at four degrees, and processed about 12 hours after collection. The samples were enriched for viral RNA as previously described [28]. About 100ml sample was filtered through 2 glass fiber and 0.2 µM PVDF filters (Millipore, cat# AP2007500 and S2GVU02RE). Of this filtrate, 60 ml was transferred to a 10 kDa cutoff centricon unit, that was previously pre-conditioned with 50 ml ultrapure water and centrifuged with 3000 g for 15 minutes at 4 °C. After centrifugation of the samples for 30 minutes at 4 °C and again 3000 g, the unit was inverted and about 400 µl concentrate was collected by centrifugation for 1000g at 4 °C for 3 minutes. The concentrate was mixed with 3 volumes of Trizol LS (ThermoFisher cat# 10296-010), and the RNA extracted using the Direct-zol RNA miniprep kit (Zymo cat# R2052) including the DNase treatment and elution with 50 µl ultrapure water according to the manufacturer’s instruction. Absence of PCR inhibitors was confirmed by mixing the sample 1:1 with total RNA from human cells followed by amplification of a human transcript. not detectable in waste water alone by RT-qPCR.

#### Reverse transcription / quantitative polymerase chain reaction (RT-qPCR)

The extracted RNA was amplified using the LunaScript reverse transcription mix (NEB cat# E3010L), with 16 µl RNA and 4 µl reaction master mix according to the manufacturer’s instructions, except for a 20 minutes incubation at 55 °C instead of 10 minutes. Afterwards, the cDNA was diluted 1:10 with ultrapure water, and 3.75 µl diluted cDNA used per qPCR reaction, using a SYBR green master mix (ThermoFisher cat# 43-643-46), and final concentrations of 250 nM of the primers on Supplementary Table S12. The presence of the proper amplicon was verified using a 2.5% TAE agarose gel.

#### ARTIC-seq of the SARS-CoV-2 genome

Amplicon sequencing libraries of the SARS-CoV-2 genome were generated using the ARTIC protocol v3 (phase I) and modified version of ARTIC protocol v4 (Phase II) [15], using 6 µl of the cDNA generated as described above as an input. The primer pools were obtained from IDT. Amplicon libraries were sequenced on an Illumina Miseq or Novaseq device with 2×250 paired-end sequencing and 20% phiX spike-in. The modified ARTIC v4 primer can be found in Supplementary Table S7.

### Berlin wastewater samples processing for very-short-reads

In order to test if the pipeline would perform reliably under industry conditions we also used it with so-called production data from the *amedes* analytical company. The sequencing was performed as follows:

45 ml of raw wastewater was centrifuged for 10 min with 10,000 x g at 4 °C. Subsequently, the supernatant was prefiltered using Filtropur S 0.45 µm filter units (Sarstedt, Darmstadt, Germany), further transferred to 100 kDa cutoff Amicon Ultra-15 units (PLHK Ultracel-PL Membran, 100 kDa Centrifugation units; Merck Sigma Aldrich Chemie GmbH, Taufkirchen, Germany) and processed according to the manufacturer’s manual.

Automated RNA isolation was accomplished using a Qia-Cube HT Extractor using the QIAamp 96 DNA QIAcube HT Kit according to the manufacturer’s protocol (Qiagen, Hildesheim, Germany).

Library preparation for NGS sequencing was performed following the complete Illumina SARS-CoV-2 sequencing workflow (Illumina COVIDSeq Test, Illumina, San Diego, USA) including RNA-to-cDNA conversion and SARS-CoV-2 targeted PCR using the ARTIC V3 primer set. The generated libraries were analyzed using NextSeq 550 and 550Dx sequencers with NextSeq 500/550 High Output Kits (v2.5; Illumina #20024906) generating 2x 37 bp paired-end output.

### Computational Methods

#### General Pipeline description

In the first step, the pipeline takes the raw reads and the additional information about used primers and adapters to perform extensive quality control. Primer trimming is done with *iVAR* [29], and *fastp* [30] is used for adapter trimming and filtering. In order to make the read quality process comprehensible, *fastQC* reports are generated after each step and summarized with additional *MultiQC* reports. The processed reads are aligned to the reference genome by *BWA Mem* [31] and various coverage statistics are taken by *SAMtools coverage/bedcov* [32]. The alignment is used further for single nucleotide variant (SNV) calling using *LoFreq* [33]. For predicting the lineage abundances, a deconvolution matrix is generated by matching the set of mutations called by *LoFreq* against the provided mutation table. The SNVs are translated to protein mutations by *Ensemble VEP* [34]. *Kraken2* [35] is used to get taxonomic classification of the unaligned reads as an additional quality measure and further insight in the samples. The mutations were filtered for a minimum read coverage, then a deconvolution method was used to calculate the proportion of lineages (more details in the section *Deconvolution analysis*) for each sample. For summarizing and visualizing the deconvolution results as a time series, by default, samples with SARS-CoV-2 reference genome coverage below 90% are discarded. For each mutation, linear regression models are used (more details in the section *Regression analysis for mutations*) to detect if any mutation is significantly increasing over time. Here discarded samples were also not included.

For each sample a set of four reports (multiQC, general QC report, taxonomic classification report, lineage report) is generated using *Rmarkdown* and *knitr*. The R-package of *plotly* is used for generating interactive visualizations. The relevant results across all provided samples are summarized by an extra report that provides insightful visualizations and accessible navigation linking to all the single reports. In this way the pipeline output provides an easily accessible overview about lineage and mutation dynamics in a communicable format but also enables extensive data exploration and access to sample-wise tables and summaries without the need for running extra scripts. PiGx SARS-CoV-2 uses snakemake [36] to define and run the workflow.

#### Deconvolution analysis

##### Model description

With **m** being a system of linear equations built by using **B** being a signature matrix constructed from the signature mutations provided as input and **f** being the proportions for the lineages the deconvolution approach can be represented as **m = f** x **B**. Similar to what has been shown before for deconvolution of cell types from gene expression profiles or methylation profiles [37], we follow the assumption that the frequency of signature mutations corresponds with the frequency of the actual lineage which is characterized by it. The difference in our approach is that we use sequence mutations and apply weights to the signature matrix in order to get more realistic prediction results.

##### Signature matrix construction

The signature matrix is obtained by matching the set of mutations found in the sample against the set of signature mutations provided as input. In case the mutation table contains mutations that are shared between lineages, it is possible that multiple lineages cannot be distinguished from each other. In this case, the signature matrix will be deduplicated leaving only one column of the duplicated lineages which will be renamed with the grouped names of all lineages showing this duplicated signature mutation “pattern”.

To make the matrix more robust, additional “reference mutations” are added as well as a reference column denoted as “Others”. Bulk frequencies for the “reference mutations” are the difference between 1 and the value of the related signature mutation.

We propose the assumption that the more signature mutations can be found for a specific lineage the higher the probability that this lineage is present with a higher proportion within the sample. We therefore weigh the signature matrix (without the reference mutations) for each lineage with the proportion of signature mutations that has been found for each specific lineage from the total number of signature mutations that was given to characterize it. For “Others” the weight was calculated proportionally to the number of mutations of the mutation table that were found. Applying weights results in less variation and more accurate predictions.

##### Regression

To deconvolute the lineage abundance we performed robust regression analysis on the signature matrix and the bulk frequency values of the signature mutations using the “Robust Fitting of Linear Models” - rlm() function from the R library MASS [38] (default settings, maxit = 100). Similar to the deconvolution method CIBERSORT [37], we set negative coefficients to 0 and normalized all coefficients to add up to 1, which then form the output value providing the predicted lineage frequency values for the provided lineages and an additional “Others” estimation.

PCR bias as well as the number of detected signature mutations influence the robustness of the results. We therefore added the additional constraint to only perform the deconvolution analysis on samples matching a minimum quality score.

##### Dealing with indistinguishable variants

After deconvolution, grouped indistinguishable lineages have to be split again. There are three possible outcomes for those groups:

Firstly, when no signature mutations for a lineage could be found, the group includes the “Others” column and is in fact “Others” only. So the grouped lineages are getting the proportion value 0, “Others” gets the deconvoluted value. Secondly, the grouped lineages are deconvoluted to 0. In this case both lineages are assigned with the value 0. Thirdly, the grouped lineages are not equal to “Others” and are getting a deconvolution value above 0. In this case the assumption for normal distribution of the lineage abundances is applied and the deconvolution value is divided by the number of grouped lineages. Each lineage is assigned this adjusted value.

#### Regression analysis for mutations time-series

For the regression analysis on mutation frequencies we applied a linear regression model using the “Fitting Linear Models” - lm - function of *R base*. The test was only performed on mutations if N(x>0) > 5 being the number of frequency values x that are above 0 across all samples. To get only increasing trends, the coefficient values were filtered for values x > 0 only. P-values were calculated by the lm-function using t-test and were filtered for p < 0.05. We report the mutation trend analysis together with and sorted by the regression coefficient as a comparable value for unstandardized effect size.

#### Pooling of samples for time series analysis and plots

For summarizing across daytime and location, the lineage frequencies are pooled by calculating the weighted average using the total number of reads of each sample as weights. The mutation frequencies are pooled by using the simple mean (without removing missing values). Figures and deconvolution plots are done with *ggplot2* [39]. For the cross-correlation analysis samples were pooled by week and the pooled unique set of non-signature mutations was counted.

#### Sample scoring for quality check

For reference genome coverage quality control, the pipeline uses the *BEDtools coverage* [40], using a BED file with the tracked signature mutation sites as input. For the regression analysis and time series plots only samples are taken in account that cover more than 90% SARS-CoV-2 genome (except for the NYC dataset).

#### *In silico* data simulation

In order to qualify and test the accuracy of the pipeline under industrial sequencing parameters, an artificial dataset containing only short single-end sequencing was simulated. The simulated dataset was generated in-silico using full genomes of 6 SARS-COV-2 lineages obtained from GISAID [41]. The genomes were used to simulate Illumina sequencing using InSilicoSeq [42] and Seqtk [43] was used to trim sequences down to 40bp of length and subsample reads. A total of 100.000 reads was generated using the following proportions: 10% P1 (gamma), 10% B.1.1.7 (alpha), 10% B.1.621 (mu), 50% C.37 (lambda), 15% Delta (B.1.617.2) and 5% B.1.1.529 (omicron).

The data was processed without primer trimming and without an additional filter for read coverage.

Accessions from the genomes used to simulate sequencing can be found on Table 1.

**Table 1:**
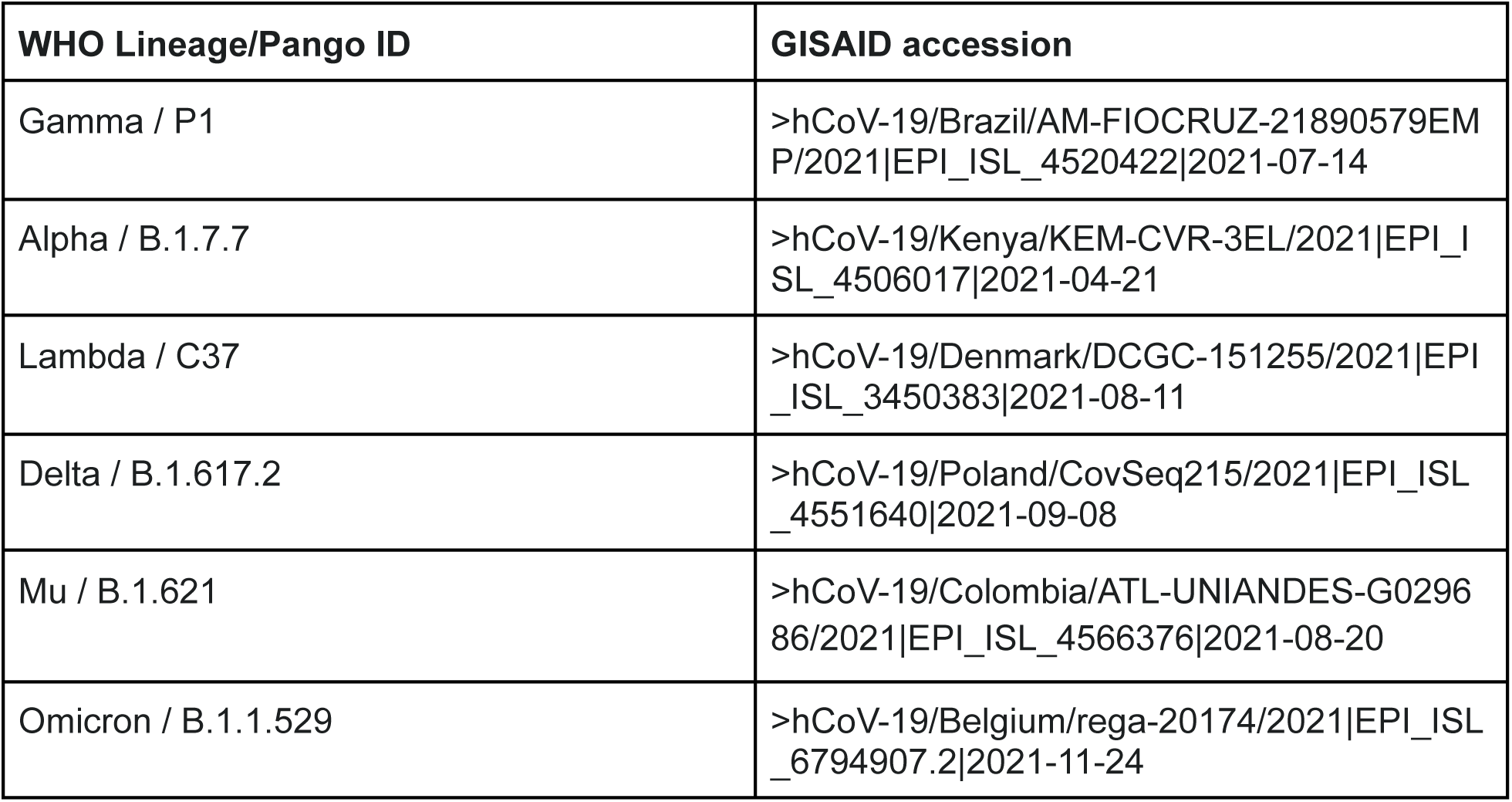
SARS-CoV-2 genomes used for *in silico* simulations.

#### Spike-in samples data acquisition

Spike-in sequencing bam files were generated by Karthikeyan et. al. [14] Data was downloaded from: https://console.cloud.google.com/storage/browser/search-reference_data. The data was processed without primer trimming and but the filter for minimal read coverage was set to 100

#### Processing of wastewater data from New York City

The data was downloaded from the Sequence Read Archive SBI with the accession number # PRJNA715712. The MiSeq data was processed with primer trimming, using the primer sequences published by the authors. The iSeq data was processed without primer trimming. For both datasets a minimal read coverage filter of 100 was applied. No genomic coverage percent cutoff was used for those datasets.

#### Data/Code Availability

The pipeline can be installed with GNU Guix and is executed with the command [ pigx-sars-cov2-ww -s {sample_sheet} {settings_file} ]. A cloud version is also being developed. Information about other alternatives like building from source or potentially a Docker image can be found on the repository. We recommend installing the pipeline with GNU Guix for its reproducibility guarantees [44]. For installation advice, documentation and code please visit the pipeline’s repository: https://github.com/BIMSBbioinfo/pigx_sars-cov-2.

#### Reproducible environment

The presented results were produced using PiGx SARS-CoV-2 version 0.0.5.

- dataset-Berlin250, dataset-NYC(RBD) (MiSeq data and all samples merged) - commit 524ed4832a6972fd695c0eeec25264188710a143
- dataset-Berlin35, dataset-NYC(RBD) (iSeq data), insilico-simulation - commit 0a150c4bec58a5a8296c870586e225e49ee2b6f8
- UCSD-spike in - commit bd87e7f2d83317e9d83f6fd81abb631af95476f6

The repository also contains the Guix manifest for this analysis (commit 4ded8c5bdc755391360e5695003d6d4085110d08). The channels file to reproduce the environment that was used for the analysis can be found in Supplementary Table S8.

#### Data access

The raw sequencing read data from Berlin wastewater samples is deposited to the Sequence Read Archive (SRA) available using the accession number #PRJNA827160.

The interactive reports that were used and produced for this pipeline can be found here:

- dataset-Berlin250 - https://bimsbstatic.mdc-berlin.de/akalin/AAkalin_pathogenomics/sarscov2_ww_reports/220225_dataset_Berlin250/index.html
- dataset-Berlin35 - https://bimsbstatic.mdc-berlin.de/akalin/AAkalin_pathogenomics/sarscov2_ww_reports/220310_dataset_Berlin35/index.html
- dataset-NYC(RBD) - https://bimsbstatic.mdc-berlin.de/akalin/AAkalin_pathogenomics/sarscov2_ww_reports/220225_dataset_NYC_RBD/index.html
- UCSD-spike in - https://bimsbstatic.mdc-berlin.de/akalin/AAkalin_pathogenomics/sarscov2_ww_reports/220309_ucsd_spikeIn/index.html
- Insilico-simulation- https://bimsbstatic.mdc-berlin.de/akalin/AAkalin_pathogenomics/sarscov2_ww_reports/220310_insilico_simulation/insilico_simulation.html

## Supporting information

SupplementaryTable_S2.2_mutations_found_Berlin35

SupplementaryTable_S2.3_mutations_found_ucsd

## Data Availability

Data is available within the links provided in the manuscript

https://github.com/BIMSBbioinfo/pigx_sarscov2_ww

https://bimsbstatic.mdc-berlin.de/akalin/AAkalin_pathogenomics/sarscov2_ww_reports/220225_dataset_Berlin250/index.html

https://bimsbstatic.mdc-berlin.de/akalin/AAkalin_pathogenomics/sarscov2_ww_reports/220310_dataset_Berlin35/index.html

https://bimsbstatic.mdc-berlin.de/akalin/AAkalin_pathogenomics/sarscov2_ww_reports/220225_dataset_NYC_RBD/index.html

https://bimsbstatic.mdc-berlin.de/akalin/AAkalin_pathogenomics/sarscov2_ww_reports/220309_ucsd_spikeIn/index.html

https://bimsbstatic.mdc-berlin.de/akalin/AAkalin_pathogenomics/sarscov2_ww_reports/220310_insilico_simulation/insilico_simulation.html

## Acknowledgements

We thank Mrs. Burzyk, Cytner, Darre, Göldner, Grunow, Heinig, Horn, Kapczynski, Klawonn, Koch, Krug, Meyer, Neideck, Schmidt, Schwarzenberg, Stroede, Zühlsdorff, and Messrs. Armbrecht, Dombrowski, Frankenstein, Halatta, Hambarsomian, Linnek, Muss, Flatau, of the Berliner Wasserbetriebe for sampling and logistic support; as well as Dr. Selinka of the Umweltbundesamt and also Mrs. Schumacher for helpful discussions. Also, we would like to thank Friederike Dündar for consultation on best practices for visualization techniques and Jonas Freimuth for code discussions and support with code development.

